# MRISeqClassifier: A Deep Learning Toolkit for Precise MRI Sequence Classification

**DOI:** 10.1101/2024.09.19.24313976

**Authors:** Jinqian Pan, Qi Chen, Chengkun Sun, Renjie Liang, Jiang Bian, Jie Xu

**Affiliations:** Department of Health Outcomes & Biomedical Informatics, University of Florida, Gainesville, FL, USA; The 2nd Affiliated Hospital and Yuying Children’s Hospital of WMU, Wenzhou, Zhejiang, China

## Abstract

Magnetic Resonance Imaging (MRI) is a crucial diagnostic tool in medicine, widely used to detect and assess various health conditions. Different MRI sequences, such as T1-weighted, T2-weighted, and FLAIR, serve distinct roles by highlighting different tissue characteristics and contrasts. However, distinguishing them based solely on the description file is currently impossible due to confusing or incorrect annotations. Additionally, there is a notable lack of effective tools to differentiate these sequences. In response, we developed a deep learning-based toolkit tailored for small, unrefined MRI datasets. This toolkit enables precise sequence classification and delivers performance comparable to systems trained on large, meticulously curated datasets. Utilizing lightweight model architectures and incorporating a voting ensemble method, the toolkit enhances accuracy and stability. It achieves a 99% accuracy rate using only 10% of the data typically required in other research. The code is available at https://github.com/JinqianPan/MRISeqClassifier.

## Introduction

Magnetic Resonance Imaging (MRI) is a fundamental and powerful diagnostic tool in the medical field, widely employed for detecting and evaluating a wide range of health conditions. Various MRI sequences, such as T1-Weighted Imaging (T1WI), T2-Weighted Imaging (T2WI), and Diffusion-Weighted Imaging (DWI), are tailored to aid in diagnosing specific conditions by highlighting different tissue properties. For instance, T1WI is highly sensitive for detecting lesions such as cysts and is commonly used to diagnose diseases affecting the brain, bones, and abdomen. ^1^ T2WI excels at revealing high water signals, making it ideal for identifying oedema and changes in water content around lesions; it is frequently used for diagnosing tumors, white matter lesions, and meningiomas. ^2^ DWI, which measures the diffusion of water molecules within tissues, is crucial in diagnosing conditions like stroke and brain tumors, as it can detect minute infarcts or edemas in the brain. ^3^

However, the introduction of new imaging sequences and the consequent rapid increase in MRI data volumes present substantial challenges, notably in data storage and image annotation. ^4^ The inconsistency in sequence naming by different manufacturers and the lack of uniform annotation standards at various imaging centers significantly complicate data management. ^5,6^ This complexity is further amplified by the varied nomenclature used by manufacturers and imaging centers, which typically remains internal and is not shared beyond the development teams. ^7,8^ Furthermore, while medical images are commonly stored in Digital Imaging and Communications in Medicine (DICOM) and Neuroimaging Informatics Technology Initiative (NIfTI) formats, the variability in labeled parameters across different institutions complicates cross-institutional data integration and processing. ^9^ Studies have also shown that up to 16% of inspected DICOM headers contain error messages, further compromising data reliability and usability. ^6,10–12^ In addition, the process of labeling medical images is not only labor-intensive but also requires highly trained personnel, creating a significant bottleneck in aggregating large image collections and severely hindering research progress. This underscores the urgent need for more streamlined annotation practices and standards in medical imaging.

Despite several studies on MRI sequence classification, research in this area remains relatively limited. For instance, Baumgartner et al. ^6^ focused exclusively on the classification of prostate MRI sequences, while Zhu et al. ^13^ and Kim et al. ^12^ concentrated on abdominal MRI sequences. Additionally, studies by Sugimori et al. ^14^, Noguchi et al. ^15^, Pablo et al. ^16^, and Ranjbar et al. ^8^ were restricted to axial plane MRI imaging of the brain. While these studies offer valuable insights into MRI classification, they typically depend on large, meticulously curated training sets composed of high-quality, waste-free images. The creation of such high-quality training datasets necessitates significant human resources for precise labeling, which is not only costly but also labor-intensive. This requirement for extensive manual effort, while beneficial for enhancing classification accuracy, may not be feasible for research teams with limited resources or for practical applications aiming to rapidly expand their scope. Such constraints highlight the need for more efficient and scalable approaches to MRI sequence classification.

To overcome these limitations, we implemented a small-sample training approach, constructing our dataset with just 1,200 images—merely one-tenth the size typically utilized in similar research. Instead of relying solely on axial plane MRI images, as is conventionally done, we incorporated three distinct views: axial, sagittal, and coronal. This adaptation better accommodates the varied MRI preservation practices observed across different hospitals. Our multi-view strategy not only streamlines the dataset construction but also enhances the model’s generalization capabilities and helps prevent overfitting. From a technical perspective, we developed an automated toolkit in Python that leverages Convolutional Neural Networks (CNNs) and voting ensemble methods. This toolkit efficiently processes and utilizes large volumes of unlabeled medical image data with minimal intervention from specialized physicians. We have made the complete toolkit code publicly available on GitHub to facilitate easy reproduction of our results by other researchers and its application to further studies. Using this toolkit, we have successfully processed and classified over 70,000 MRI data from the National Alzheimer’s Coordinating Center (NACC) ^17^, generating a structured, high-quality dataset. These datasets not only serve as a valuable resource for medical research but also underscore the significant potential of small-sample learning methods in practical applications.

## Methods

### Data Source

Our study utilized the data from NACC, which maintains one of the largest and most comprehensive Alzheimer’s datasets. ^17^ Established over the past two decades, this dataset has been developed in collaboration with more than 42 Alzheimer’s Disease Research Centers (ADRCs) across the United States. The MRI data from NACC, originating from a mixed protocol, can best be described as a convenience sample of images that were voluntarily submitted by several ADRCs. Based on the structural data provided by NACC, we obtained a total of 73,449 valid image data files, which included both MRI data and associated structural information.

The MRI images in the NACC dataset were classified into six categories based on the MRI sequences: T1WI, T2WI, FLAIR, DWI, DTI, and other images. The “other” category encompasses all sequences not fitting the previous five classifications according to NACC. Examples of middle slices from these six MRI sequence types are shown in **Figure 1**. However, the labeling accuracy from NACC is suboptimal. Some files do not conform to any specified categories, or the image data may only include localizers from MRI scans, or even contain discarded MRI images. Our study aimed to accurately label MRI sequences and reclassifying any unwanted sequences and scrap images as “other.”

**Figure 1.**
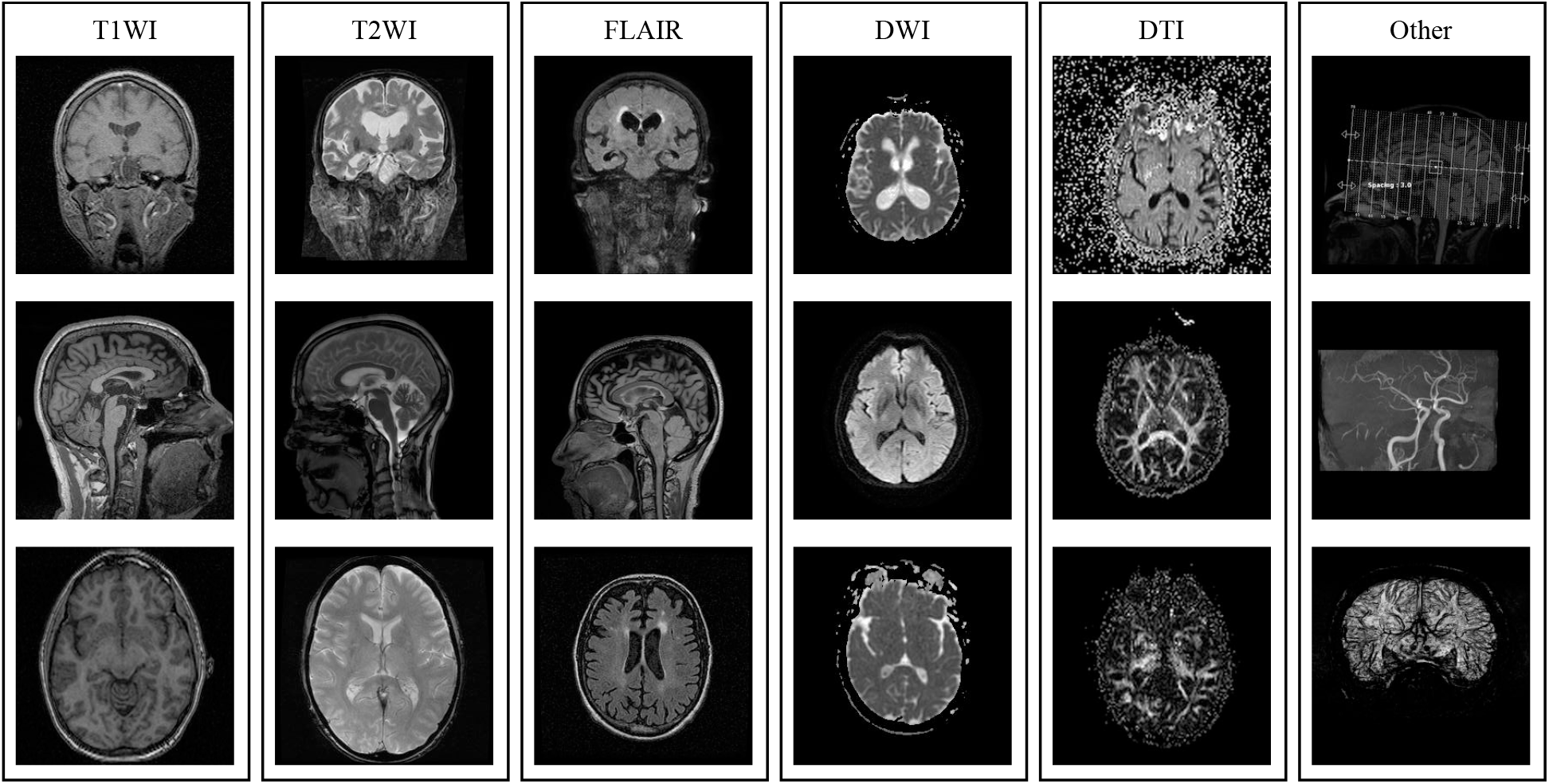
Examples of <u>middle</u> slices from six MRI sequences types: T1-Weighted Imaging (T1WI), T2-Weighted Imaging (T2WI), Fluid Attenuated Inversion Recovery (FLAIR), Diffusion-Weighted Imaging (DWI), Diffusion Tensor Imaging (DTI), and others.

### Data Preprocessing

The initial challenge we encountered involves processing a vast dataset containing 4 TB of NIfTI image files and JSON header files, spread across thousands of compressed packages. To enhance processing efficiency, we first converted all .nii files to .nii.gz format and reorganized the file structure accordingly. Concurrently, we extracted metadata from the JSON files and saved it into CSV files. This preprocessing step enabled us to compress the dataset to a more manageable size of 656 GB.

The MRI data were originally collected in 3D volumes, with varying contrast levels between proximal and middle slices. We specifically targeted the first proximal slices and middle slices due to their contracting characteristics. ^15^ We processed this data along the third axis, extracting these specific slices and converting them into JPG format to create two distinct 2D datasets. In contrast to approaches like AINNAR ^15^, which focus on specific 2D views such as axial, coronal, or sagittal, we opted for a more inclusive strategy. By avoiding the targeting of specific 2D views, we minimized the risk of overfitting our CNN models to particular slice orientations.

Given that both datasets originated from the same 3D dataset, we were able to apply the same set of labels to each. We used the”SeriesDescription” field from the JSON metadata to guide the initial categorization of the images. To ensure accurate labeling, we randomly selected 200 images from each category of the middle slices, which were then manually annotated by a radiologist. After the labeling process and necessary adjustments, we compiled a final dataset of 1,200 images, with each category represented by 200 images.

### Model

In this study, we employed eight fundamental CNN architectures across nine different variants: AlexNet ^18^, GoogLeNet ^19^, ResNet18^20^, DenseNet121^21^, EfficientNet B0 and EfficientNet V2 Small^22^, ConvNeXt Tiny ^23^, MobileNet V3 Small ^24^, and VGG11^25^.

#### AlexNet^18^

This seminal CNN model, comprising five convolutional layers and three fully-connected layers, marked a significant milestone in deep learning. It set a new standard for image recognition tasks and catalyzed advancements in the field.

#### GoogLeNet^19^

Introduced with the innovative “Inception module”, GoogLeNet enhances parameter efficiency and network depth. Its architecture has been pivotal in advancing the capabilities of image recognition and classification systems.

#### ResNet18^20^

Part of the residual network family, this lightweight model has 18 layers and uses residual connections to address gradient vanishing in deep neural networks. This allows for learning complex features through deeper network structures, making it a popular choice for image classification and other computer vision tasks.

#### DenseNet121^21^

This model, part of the DenseNet series, includes 121 layers. It is characterized by direct connections from each layer to all preceding layers, promoting feature reuse and reducing the model’s parameter count.

#### EfficientNet B0 and EfficientNet V2 Small ^22^

EfficientNet B0, the series’ initial model, systematically scales network depth, width, and resolution for optimal performance. EfficientNet V2 Small, designed for faster training and greater efficiency, improves upon the original architecture by introducing features that enhance training friendliness and maintain or boost performance while reducing parameters.

#### ConvNeXt Tiny^23^

A modern take on traditional CNN structures, it incorporates layer normalization and redesigned convolutional layers similar to those in Transformers. This design enhances computational efficiency and performance, particularly for image classification and other visual tasks on resource-limited devices.

#### MobileNet V3 Small ^24^

An efficient, lightweight deep learning model optimized for edge devices and mobile applications, it uses hardware-aware Neural Architecture Search (NAS) to optimize its structure and combines h-swish and SE (Squeeze-and-Excitation) modules to boost performance.

#### VGG11^25^

A simpler model in the VGG series, it consists of 11 layers, including 8 convolutional layers and 3 fully-connected layers. Known for its uniform architecture and use of small 3 × 3 convolutional kernels, VGG11 excels in capturing fine image details and is widely used in image classification and recognition tasks.

For our analysis, we combined the predictions from these nine different models using a voting ensemble method. ^26^ This technique aggregates predictions from multiple models, with the plurality vote determining the final output. By leveraging this ensemble approach, we aimed to minimize the likelihood of errors from any single model and enhance overall prediction accuracy.

### Experiments

As shown in **Figure 2**, we employed a 10-fold cross-validation strategy along with stratified sampling to ensure balanced representation of all categories in each fold. This method partitioned the training data into ten distinct subsets, maintaining proportional representation of each class. Each fold was independently used to train and validate the models, ensuring that every data point contributed to both training and validation phases. After 100 epochs of training per fold, we selected the nine best-performing models based on validation accuracy. These models were subsequently utilized to make predictions on the test set. The predictions from these models were aggregated to evaluate individual mode accuracy and were further combined using a voting ensemble method, wherein predictions from multiple models were integrated to form a consensus output for the final prediction.

**Figure 2.**
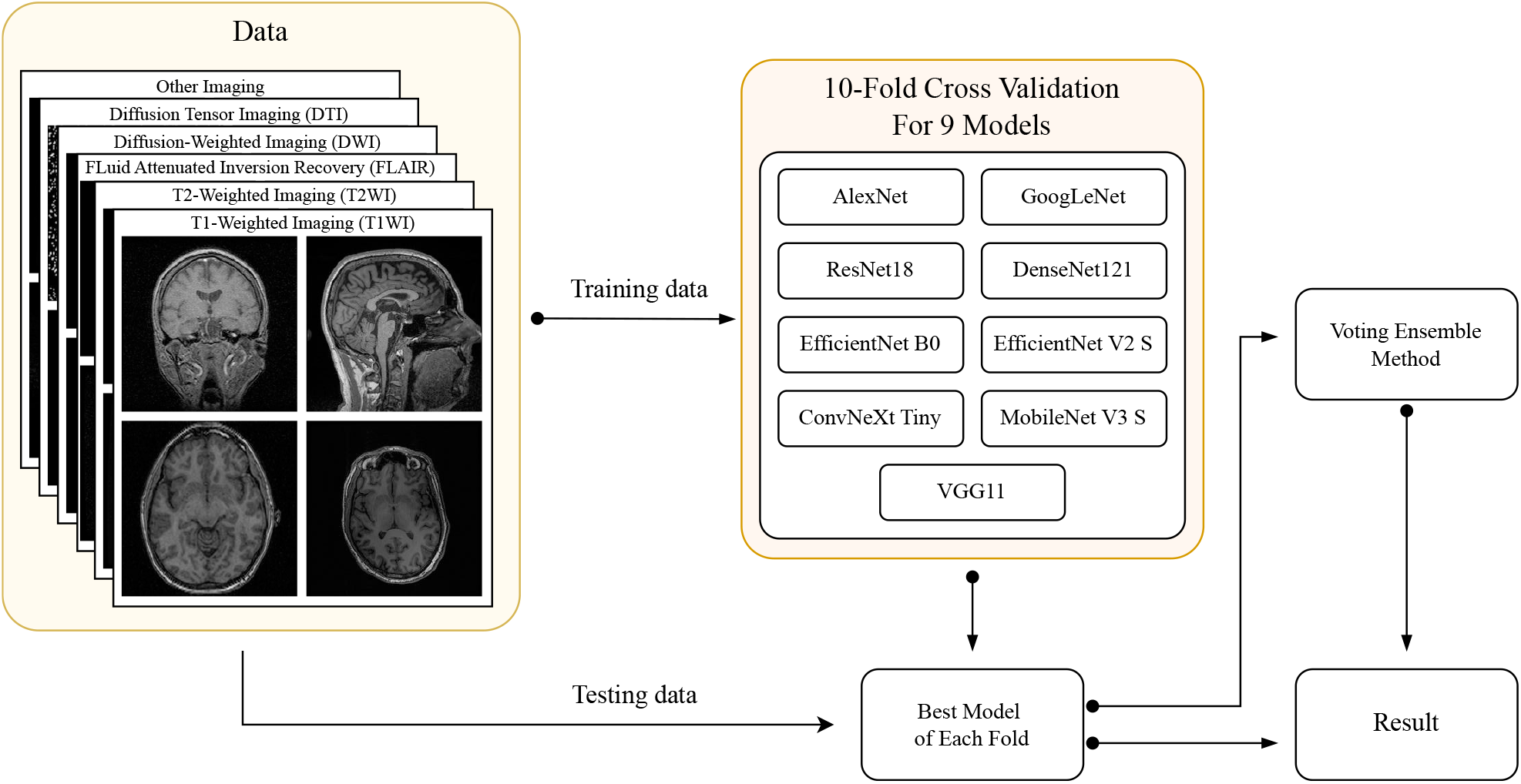
MRI Sequences Classification Pipeline: (1) The training data is distributed among nine different models for 10-fold cross-validation. (2) The best-performing models from each validation fold are selected and applied to the test data to obtain predictions. (3) A voting ensemble method combines the predictions from all nine models to produce the final result.

All models in our study were implemented in PyTorch^27^ and trained on a single NVIDIA A100 GPU (equipped with 8 GB of RAM) and one CPU core. We initialized the models using pre-trained weights from the ImageNet dataset^28^, which significantly improved both the training efficiency and final performance. For optimization, we used the Stochastic Gradient Descent (SGD) algorithm, setting an initial learning rate of 0.001 with a momentum of 0.9. The datasets were randomly split using stratified sampling into an 80%-20% ratio for training and testing, with 10-fold cross-validation consistently applied throughout the model training and validation phases. Specifically, the training dataset consisted of 960 images, with 160 images per category, while the test dataset comprised 240 images, with 40 images for each category. A batch size of four was used. To enhance model generalization, data augmentation techniques such as random horizontal flipping and resizing images to 224 × 224 pixels were applied to match the models’ input requirements. We also used normalization parameters consistent with those of the ImageNet dataset. Cross Entropy Loss was utilized as the loss function, with accuracy being the primary metric for evaluating model performance, highlighting our emphasis on correctly classifying images.

## Result

**Table 1** summarizes the results, including the mean accuracy and standard deviation, from a 10-fold cross-validation on both datasets. Among all the basic models evaluated, EfficientNet V2 Small stood out with its performance. It achieved the highest accuracy on the proximal first slice dataset at 93.17%, outperforming the lowest-rated model by 7.83% and exceeding the second-best model by 0.91%. On the middle slice dataset, it ranked as the top-performing model with an accuracy of 99.17%, which is 2.75% higher than the worst-performing model and 0.29% higher than the model in second place. DenseNet121 secured the third position on the proximal first slice dataset with an accuracy of 92.17%, and closely followed the leader on the middle slice dataset with a 98.79% accuracy, just 0.38% less. AlexNet lagged behind on both datasets, recording the lowest accuracies.

**Table 1.** Accuracy (%) ± standard deviation of different models on the proximal first slice and middle slice, evaluated using 10-fold cross-validation.

| Models | Promixal First Slice Dataset | Middle Slice Dataset |
| --- | --- | --- |
| AlexNet | 84.42 ± 1.55 | 96.42 ± 0.71 |
| GoogleNet | 91.42 ± 0.66 | 98.88 ± 0.34 |
| ResNet18 | 90.96 ± 2.15 | 98.83 ± 0.55 |
| DenseNet121 | 92.17 ± 1.21 | 98.79 ± 0.63 |
| EfficientNet B0 | 92.25 ± 1.32 | 98.79 ± 0.41 |
| ConvNeXt Tiny | 89.33 ± 1.52 | 98.33 ± 0.52 |
| EfficientNet V2 Small | 93.17 ± 0.97 | 99.17 ± 0.34 |
| MobileNet V3 Small | 90.46 ± 1.41 | 98.00 ± 0.94 |
| VGG11 | 90.21 ± 1.50 | 98.12 ± 0.63 |
| Voting Model | 94.62 ± 0.84 | 99.38 ± 0.22 |

The application of the voting ensemble method further improved the results. It surpassed the runner-up by 0.45% with an accuracy of 94.62% on the proximal first slice dataset and edged out the second place by 0.17% with an accuracy of 99.38% on the middle slice dataset. Moreover, the stability of the model optimized with the voting ensemble method was outstanding, exhibiting the lowest standard deviation among almost all the models.

We also presented the confusion matrices of the two most representative folds from each dataset (**Figure 3**). In the middle slice dataset, most misclassifications primarily involved incorrectly classifying the “other” category as T1WI. In the first proximal slice dataset, in addition to the types of misclassification observed in the middle slice dataset, there was additional confusion among T1WI, T2WI, and FLAIR.

**Figure 3.**
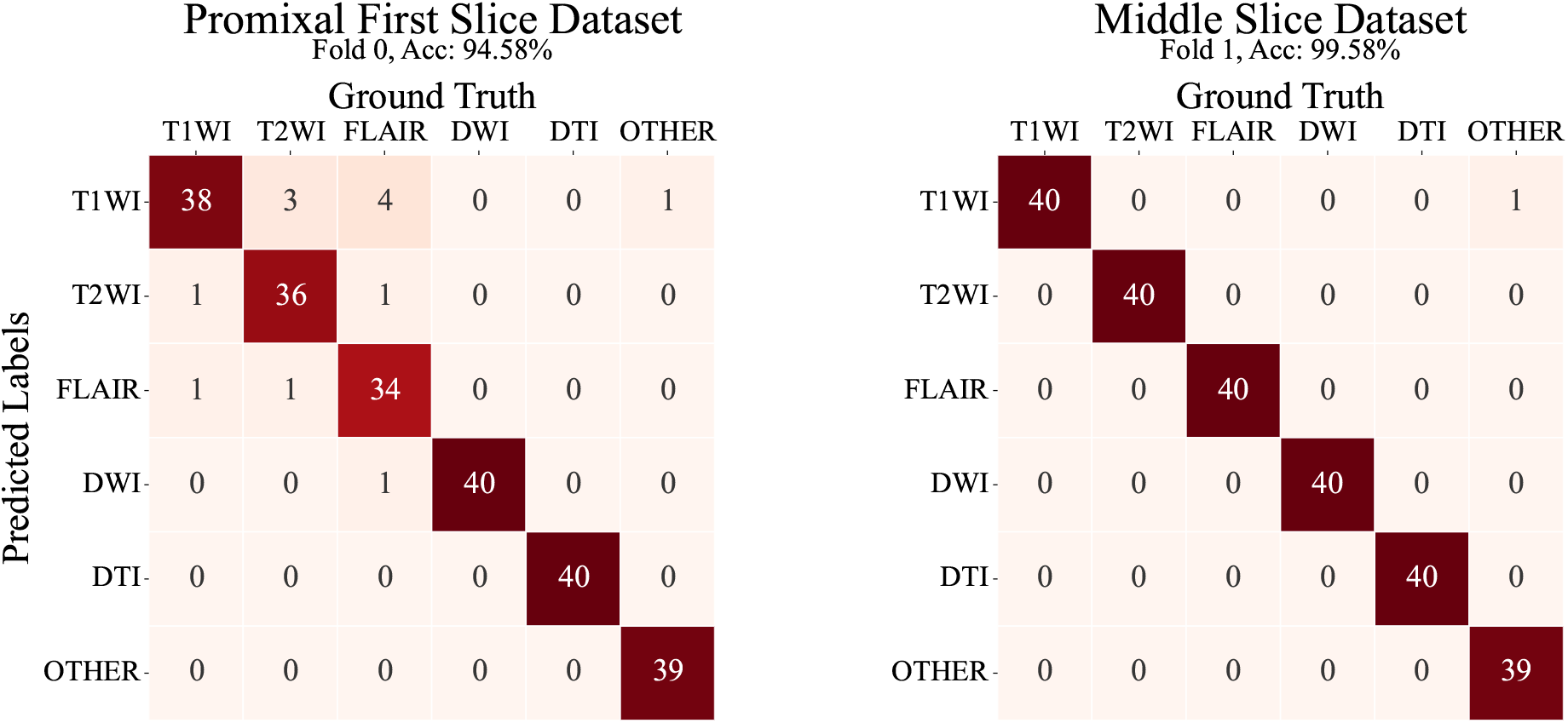
Confusion matrices of predicted MRI image types across different datasets, showcasing an accuracy of 94.58% for the Proximal First Slice Dataset (Fold 0) and 99.58% for the Middle Slice Dataset (Fold 1). Each type consists of 40 images, with matrix values representing the count of each type based on ground truth (manually annotated by a radiologist) and predicted labels.

## Discussion

Our results indicate that EfficientNet V2 Small and DenseNet121 outperform the other seven models in small-sample classification tasks. The superior performance of EfficientNet V2 Small can be attributed to its auto-scaling capabilities and the optimization of both training speed and parameter efficiency. Meanwhile, DenseNet121 excels due to its feature reuse mechanism, making both models particularly effective for handling small-sample datasets. In contrast, AlexNet model struggled with small samples datasets due to its simpler structure and lack of modern enhancements such as residual or dense connections. Moreover, AlexNet’s large number of parameters tends to induce overfitting. Contemporary models like MobileNet and EfficientNet mitigate overfitting risks more effectively by optimizing network structures and parameters, thus enhancing both performance and resource efficiency.

The adoption of voting ensemble methods further improved performance by aggregating predictions from multiple models. This method not only increased the stability and robustness of our model but also mitigated the risks of random errors or overfitting, particularly with sparse data. By combining predictions through a majority-rule process, ensured reliable results even when individual models performed suboptimally. This approach also demonstrated versatility by leveraging the strengths of diverse model architectures, balancing bias and variance, and improving tolerance to errors. Compared to previous studies,^6,8,12–16^ which typically required several thousand to tens of thousands of training samples, our tool achieved comparable or superior accuracy rates with a smaller dataset. This highlights the effectiveness of our approach.

Our analysis of model performance, comparing predicted labels with ground truth, is illustrated in **Figure 4**. Selected examples from two datasets demonstrate various prediction errors. Notably, images with index numbers 141 showed errors across both datasets. In both datasets, images with index numbers 141 are identical, and their header files list the ProtocolName as “localizer”, indicating a localization scan. However, these images bear similarities to T1WI, leading to misclassification by the model. These images are labeled as discarded images, which, as previously discussed, are typically excluded from radiological diagnostics and categorized as “other.” In the middle slice dataset, image number 11 was incorrectly classified from DTI to low b-value DWI due to the subtle differences between DTI and DWI, especially when image contrast is not pronounced. Image number 206 was influenced by image number 141, resulting in misclassification. Additionally, in the proximal first slice dataset, images numbered 84 and 88 were inaccurately predicted due to poor image quality, such as only showing part of the skull, highlighting the model’s limitations with low-quality images. Moreover, the model incorrectly classified the FLAIR image with index number 115 as a DWI due to the prominent central highlight of the image, revealing a specific shortcoming in recognizing certain types of images. In the case of image index 224, which features bones, the model also made an erroneous judgment. These results indicate that although the model generally performs well, it struggles with lower quality or less informative images. Future efforts should aim to enhance the model’s discriminatory capabilities, particularly for edge cases, to boost overall prediction accuracy.

**Figure 4.**
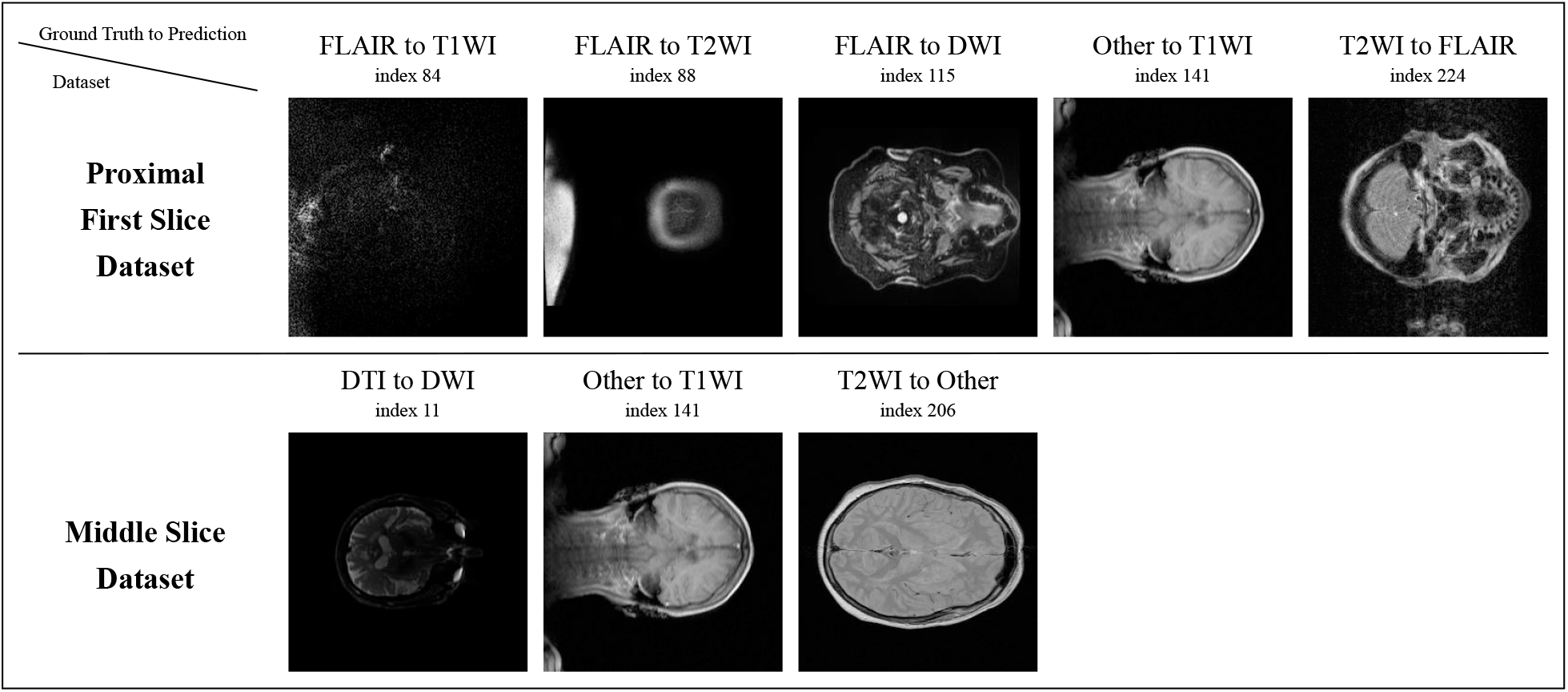
Examples of prediction errors from the test set for both datasets. Each image is labeled with the ground truth and predicted label. The index indicates the image number within test set.

Moreover, this study was exclusively trained and tested on brain MRI data, which limits the effectiveness of the developed deep learning architecture to brain MRI alone. As highlighted in the study by Kim et al.^12^, the research could expand to include different types of MRI data, such as chest and abdominal MRIs. This extension would not only validate the model’s generality in processing MRI images from various body regions but would also enhance its accuracy and practical utility in clinical applications.

In future research, we aim to enhance the training methodology and broaden the applications of our model. Initially, we observed that the model’s misclassification of T1WI, T2WI, and other MRI images when analyzing middle slice data stemmed primarily from the image quality rather than erroneous model assessments. To rectify this, we intend to refine the classification labels by dividing the “other” category into more specific subcategories, such as “other sequences”, “T1WI discarded”, “T2WI discarded”, etc. This modification will enable the model to more precisely recognize and differentiate the subtle variances among MRI images, thereby boosting overall classification accuracy. Furthermore, we plan to explore the integration of classified MRI data with other data types to develop multimodal datasets. For instance, the NACC dataset includes structured data and cerebrospinal fluid (CSF) bio-experimental data. By combining these with our MRI dataset, we aim to investigate and predict clinical outcomes such as Alzheimer’s Disease (AD), mild cognitive impairment (MCI), and the progression from MCI to AD. This multimodal approach will not only augment the model’s capacity to understand and forecast complex medical conditions but could also enhance diagnostic accuracy and support personalized treatment strategies. Through these initiatives, we hope to advance the model’s performance and extend its relevance and impact in clinical settings.

## Conclusion

In this study, we developed a deep learning toolkit tailored for MRI datasets characterized by small and unrefined samples, achieving effective and accurate classification of MRI sequences. The adoption of a voting ensemble method further improved the model’s classification accuracy and stability across various testing scenarios. Despite these advancements, our results also exposed challenges, particularly with lower-quality images or those lacking complete information, indicating areas where model performance can be further improved. Future research will focus on refining model architecture and training processes, as well as incorporating multimodal datasets to leverage a broader range of medical information, thereby deepening diagnostic capabilities.

## Data Availability

https://naccdata.org/
https://github.com/JinqianPan/MRISeqClassifier

## Acknowledge

This work was partially supported by a grant from the Ed and Ethel Moore Alzheimer’s Disease Research Program of the Florida Department of Health (FL DOH #23A09). The NACC database is funded by NIA/NIH Grant U24 AG072122. NACC data are contributed by the NIA-funded ADRCs: P30 AG062429 (PI James Brewer, MD, PhD), P30 AG066468 (PI Oscar Lopez, MD), P30 AG062421 (PI Bradley Hyman, MD, PhD), P30 AG066509 (PI Thomas Grabowski, MD), P30 AG066514 (PI Mary Sano, PhD), P30 AG066530 (PI Helena Chui, MD), P30 AG066507 (PI Marilyn Albert, PhD), P30 AG066444 (PI John Morris, MD), P30 AG066518 (PI Jeffrey Kaye, MD), P30 AG066512 (PI Thomas Wisniewski, MD), P30 AG066462 (PI Scott Small, MD), P30 AG072979 (PI David Wolk, MD), P30 AG072972 (PI Charles DeCarli, MD), P30 AG072976 (PI Andrew Saykin, PsyD), P30 AG072975 (PI David Bennett, MD), P30 AG072978 (PI Neil Kowall, MD), P30 AG072977 (PI Robert Vassar, PhD), P30 AG066519 (PI Frank LaFerla, PhD), P30 AG062677 (PI Ronald Petersen, MD, PhD), P30 AG079280 (PI Eric Reiman, MD), P30 AG062422 (PI Gil Rabinovici, MD), P30 AG066511 (PI Allan Levey, MD, PhD), P30 AG072946 (PI Linda Van Eldik, PhD), P30 AG062715 (PI Sanjay Asthana, MD, FRCP), P30 AG072973 (PI Russell Swerdlow, MD), P30 AG066506 (PI Todd Golde, MD, PhD), P30 AG066508 (PI Stephen Strittmatter, MD, PhD), P30 AG066515 (PI Victor Henderson, MD, MS), P30 AG072947 (PI Suzanne Craft, PhD), P30 AG072931 (PI Henry Paulson, MD, PhD), P30 AG066546 (PI Sudha Seshadri, MD), P20 AG068024 (PI Erik Roberson, MD, PhD), P20 AG068053 (PI Justin Miller, PhD), P20 AG068077 (PI Gary Rosenberg, MD), P20 AG068082 (PI Angela Jefferson, PhD), P30 AG072958 (PI Heather Whitson, MD), P30 AG072959 (PI James Leverenz, MD).

